# Prevalence and social determinants of breastfeeding practices in urban slums and urban non-slum areas in India: a comparative analysis

**DOI:** 10.1101/2025.04.17.25326006

**Authors:** Suliat Fehintola Akinwande, Chiamaka Okonkwo, Kalonde Malama, Thabani Nyoni

## Abstract

Understanding how the prevalence and determinants of breastfeeding practices differ between urban slum areas, where living conditions are stringent, and urban non-slum areas, which have relatively more resources is key to context-specific interventions. We conducted a comparative analysis to investigate the prevalence of breastfeeding practices in the urban slum and urban non-slum areas of India. We also used the socio-ecological framework to assess the individual, community, and policy-level correlates of breastfeeding practices. Secondary analysis of data from the National Family and Health Survey (2015-2016) in India was conducted to estimate the prevalence of early breastfeeding initiation (EBFI) and exclusive breastfeeding (EBF) of children living in urban slum and urban non-slum areas, and the prevalence estimates were further stratified by the seven states where slums were sampled. Logistic regression analysis was used to examine the correlates of breastfeeding practices. EBFI was significantly higher in the urban slum areas (50.4%) compared to urban non-slum areas (37.4%). In contrast, EBF was lower in urban slums (50.1%) than in urban non-slum areas (55.8%). EBFI varied widely across states, with urban slum areas showing higher rates in Maharashtra and Tamil Nadu, but lower in Delhi and West Bengal compared to urban non-slum areas. EBF was generally higher in urban non-slum populations, though some slum areas (e.g., Madhya Pradesh and Telangana) reported notably high rates. In urban slums, community-level factors, such as being part of Scheduled Tribes and Other Backward Classes (other marginalized classes), were associated with higher EBFI. At the policy level, home-based child delivery was associated with EBF in urban non-slum areas. The study showed that breastfeeding practices need to be urgently addressed in both the urban slum and urban non-slum areas, and the wider health determinants, such as community and policy level factors should be considered in designing effective interventions.

## Introduction

Optimal breastfeeding practices have been recommended by WHO and UNICEF as key to reducing child mortality and could save over 820,000 children yearly (1). Early breastfeeding initiation (EBFI) within an hour of birth and exclusive breastfeeding for six months is fundamental to the health and development of children (2). The breast produces colostrum, which is a special milk secreted in the first 2-3 days of childbirth, it is rich in white blood cells, antibodies, protein, minerals, and fat-soluble vitamins (A, E and K). Breastmilk contains all the required nutrients and immunity for children in the first six months of life (3). Exclusive breastfeeding helps prevent diarrhea and infections, supports intellectual and motor development, and reduces the risk of chronic diseases (3,4). Randomized control trials from low and middle-income countries showed that 72% of diarrhea admissions and 57% of respiratory infections were prevented by breastfeeding (4). Even in high-income countries, breastfeeding has been reported to prevent infectious diseases, mortality from necrotizing enterocolitis and sudden infant death syndrome (3).

More than 1 in 4 deaths of children aged under 5 years occurs in under-resourced environments such as urban slum areas, however, little is known of the breastfeeding practice of children living in these areas (5). Urban slum areas are characterized by poor sanitation, overcrowding, inadequate ventilation, lack of potable water, and rapid spread of diseases which increase the risk of stunting, wasting and infections among children (6). People living in urban slum areas are also more likely to experience food insecurity, economic insecurity, low levels of education and poor access to and utilization of healthcare, which are contextual factors associated with malnutrition (7). A meta-analysis of Demography and Health Surveys from 28 developing countries, indicated that children living in urban slum areas had a higher risk of stunting compared to children in urban non-slum areas (8). With the increasing migration trend from rural and small urban cities to urban slum areas, it is necessary to examine the feeding practices of children living in urban slum areas to meet UNICEF’s goal of ending all forms of malnutrition.

Children aged under five years in India still experience suboptimal health, growth, and survival outcomes due to malnutrition. Globally, India accounts for 669,956 under-five mortality, approximately 40 million children who have stunted growth, and 21 million who are affected by wasting (9,10). Of the 1.1 billion people in the world who live in urban slum areas, India accounts for 65.49 million people and 13.9 million households living in these under-resourced areas (11,12). Despite these statistics, the projected increase of urban slums to 105.7 million in India, and the likelihood that these poor nutritional outcomes are exacerbated in urban slums, few studies in India have assessed EBF in urban slums (13). Among the few studies assessing EBF in urban slum areas in India, application of the WHO guidelines of measuring EBF among 0 to 5-month-olds is inconsistent; only one study used the WHO age guideline, while other studies include 6-month-olds, which raises concerns about the validity of their findings (14–16). Moreover, studies are missing a comparative aspect to contrast breastfeeding practices in the urban slum and non-slum areas, which could help inform best practices for both contexts. Further, studies have mainly looked at individual-level factors (e.g., child’s birth order, mother’s age, educational level of the mother, etc.) and not addressed the wider determinants of breastfeeding practices in the community (societal class, religion, and culture) and policy (health facilities, budget allocation, housing, etc.) levels (14–16)

We designed a study to address the limited evidence base to inform context-specific interventions for nutrition programmes in urban slum and urban non-slum areas. Our study aims to:

1. Compare the prevalence of breastfeeding practices in the urban slums and urban non-slum areas in 7 states in India
2. Identify the determinants of breastfeeding practices at the individual, community, and societal levels

### Theoretical framework

Based on the socio-ecological model, individual, community and policy-level characteristics have interlinked effects on breastfeeding practices (17). The individual-level factors relate directly to the mother and child characteristics like the child’s sex, birth order, and mother’s level of education (17). Due to the prevalent patriarchal system in India, gender differences in the nutrition of children exist, with girls being the most disadvantaged (18). Specifically, the patriarchal system renders girls less likely to acquire education, and when these girls eventually become mothers, they are less likely to receive and/or understand key health information (19). Community-level factors influencing nutrition include sociocultural norms and religion (20). Studies have shown that women in India from disadvantaged strata/castes and lower socio-economic classes often experience different socio-cultural factors that influence their well-being and that of their child during childbirth, and these practices vary from region to region (21,22). Such practices include discarding colostrum and isolating mother and child at birth (23). At the policy level, slum dwellers have been historically marginalized and denied access to health services, which makes mothers in slum areas unlikely to access quality care during childbirth and postpartum (6).

## Methods

### Data source and sampling

Secondary analysis was conducted using the fourth National Family and Health Survey (NFHS) data of India (2015–2016). The Ministry of Health in India and International Institute of Population Science in Mumbai coordinated the NFHS survey, which takes place every five years. NFHS-IV sampling was designed to include urban slum areas in seven states in India (Maharashtra, Tamil Nadu, Uttar Pradesh, West Bengal, Telangana, Madhya Pradesh, and Delhi). A 3-stage stratified random sampling methodology was employed in the survey for urban areas. The first stage was the selection of the primary sampling using the 2011 census as the sampling frame, and the probability proportional to size sampling was used in the selection of primary sampling units. In the final stage, households were randomly selected based on systematic sampling from each primary sampling unit. Urban slums were identified using the following criteria: “ 1) a compact area of at least 300 population or about 60-70 households of poorly built congested tenements, in an unhygienic environment usually with inadequate infrastructure and lacking in proper sanitary and drinking water facilities’’ The following were criteria used to identify slums in NFHS-IV; “1) all specified areas notified as ‘slum’ by state/local government and union territory (UT) administration under any act; 2) all areas recognized as ‘slum’ by state/local government and UT Administration which may not have been formally notified as slum under any act; 3) a compact area of at least 300 population or about 60-70 households of poorly built congested tenements, in unhygienic environment usually with inadequate infrastructure and lacking in proper sanitary and drinking water facilities”. The different areas within the cities were classified as “slum” and “non-slum” area respectively (24). Further details on the survey sampling method have been described elsewhere (25).

### Study population

Women residing in the above mentioned states were interviewed by trained data collection staff using standardized questionnaires based on WHO format (S1 file). Information on the feeding practices of the youngest children under the age of 2 years was collected from the mothers based on 24-hour recall, except for EBFI, which was based on the mother’s response to how soon breastfeeding was initiated post-delivery.

#### Inclusion criteria

We included women aged 15 to 49 years, and also last-born children living with their mothers, to reduce the potential impacts of recall bias and erroneous information.

### Outcome variables: breastfeeding practice indicators

The breastfeeding practices were assessed using the WHO core guidelines (26). EBFI was the proportion of children born in the last 24 months who were breastfed within one hour of birth. While EBF under 6 months was the proportion of infants aged 0-5 months who were fed only with breast milk, including medications.

### Explanatory variables

The individual, community and policy-level variables were selected based on previous studies and data collected by the NFHS questionnaire (14–16,27). The individual-level variables were the child’s sex, child’s birth order (1^st^, 2^nd^-4^th^, ≥5^th^), previous birth interval (<24 and ≥ 24 months), age of mother (15-19, 20-34, and ≥35years), religion (Hinduism, Muslim or others), education level of the father and mother (no education, primary, secondary and above), household wealth index, number of antenatal clinics visits (1-2, 3-6, and ≥7), if mother listens to radio, mother reads newspapers, or mother watches television (not at all/at least once a week or almost every day). Community-level variables were class (upper class, scheduled castes, scheduled tribes, and other backward class— henceforth referred to as ‘marginalized class’, and state of residence. While the policy-level variable was the place of child delivery (health facility or non-health facility). In the survey, the household wealth index was divided into five quintiles (poorest, poorer, middle, richer, and richest) using principal component analysis, and the scores were based on the number and kinds of consumer goods owned, including television, bicycle, cars and household features like drinking water, floor material and toilet facilities (International Institute for Population Sciences and ICF, 2017).

### Ethical consideration

Our study utilized publicly available data from the NFHS 2015-2016 survey, which contains no identifiable participant information and is accessible via the Demography Health Survey (DHS) website. The Institutional Review Board of the International Institute for Population Sciences (IIPS), Mumbai, and the ICF Institutional Review Board granted ethical approval for the survey. Before each interview, written informed consent was obtained from all participants during data collection. The NFHS-IV dataset is fully anonymized, ensuring that individual respondents cannot be identified. We accessed the datasets with permission from the DHS Program on the 29^th^ of July 2022. Since the data were de-identified and accessed following established protocols, separate ethical approval was not required, and data confidentiality was maintained throughout the analysis.

### Statistical analysis

Data analysis was performed using Stata Version 15.0. We first conducted a descriptive analysis of all variables to determine frequencies and proportions. The prevalence of each breastfeeding practice (EBFI and EBF) stratified by urban slum and urban non-slum was estimated and reported with 95% confidence intervals. EBFI and EBF rates for each state were also estimated by urban slum and non-slum areas. Logistic regression was performed to examine the relationship between breastfeeding practices and individual-, community-, and policy-level characteristics. Bivariate analysis was used to determine the variables to include in the multivariate analysis model. The *p*-value <0.20 was used as the cut-off for inclusion in the final model and for the multivariable analysis; *p*-value <0.05 was considered statistically significant in the final multivariable model. The variance inflation factor was used to test for multicollinearity. Stata’s survey estimation command was used to account for complex survey weights and sampling design of DHS surveys.

## Result

### Individual-level characteristics of the participants

The sample size consisted of 933 and 2315 mother-child pairs in the urban slum and non-slum areas respectively (Table 1). The median age of the children in both areas was 11 months with an interquartile range (IQR) of 6-17 months. While the median age of mothers in the slum was 25 years old (IQR=23-28) and 26 years old (IQR=23-29) in the non-slum areas. Mothers who had secondary and higher education were more in the urban non-slums (77.3%) than mothers in the urban slums (68.7%). The majority of fathers in the slum (78.9%) and non-slum (87.7%) had no education. Over 90% of mothers in both urban slums and urban non-slums delivered babies at the health facilities. Most mothers watched television almost every day in the slum (72%) and non-slum areas (83.2%), but less than 20% watched the news or listened to the radio in both areas. In the slum areas, only 5.9% of participants belonged to the richest wealth quintile, compared to 21.1% in the non-slum areas.

**Table 1.** Individual, community and policy level characteristics of the study participants in the urban-slum and non-slum in India.

|  | Urban slum areas<br>(N=933) | Urban non-slum areas<br>(N=2315) |
| --- | --- | --- |
|  | n (%) | n (%) |
| *Individual level characteristics* |  |  |
| Sex¶ |  |  |
| →Male children¶ | 473 (50.7) | 1208 (52.1) |
| →Female children□ | 460 (49.3) | 1107 (47.8) |
| Children's age (months) ¶ |  |  |
| →<6 | 165 (17.7) | 521 (22.5) |
| →6-<24□ | 768 (82.3) | 1794 (77.5) |
| Birth order¶ |  |  |
| →1<sup>st</sup>¶ | 415 (44.4) | 950 (41.0) |
| →2<sup>nd</sup>-4<sup>th</sup>¶ | 499 (53.5) | 1301 (56.2) |
| →5<sup>th</sup> and above□ | 19 (2.1) | 64 (2.7) |
| Mother's age (years) ¶ |  |  |
| →15-19¶ | 59 (6.3) | 39 (1.7) |
| →20-34¶ | 831 (89.1) | 2123 (91.7) |
| →≥35□ | 43 (4.6) | 153 (6.6) |
| Mother's Education¶ |  |  |
| →No education¶ | 174 (18.7) | 283 (12.2) |
| →Primary school¶ | 117 (12.6) | 241 (10.4) |
| →Secondary and above □ | 640 (68.7) | 1791 (77.4) |
| Father's Education¶ |  |  |
| →No education¶ | 736 (78.9) | 2030 (87.7) |
| →Primary school¶ | 30 (3.3) | 26 (1.1) |
| →Secondary and above □ | 166 (17.8) | 259 (11.2) |
| Antenatal care visits¶ |  |  |
| →None¶ | 62 (6.6) | 153 (6.6) |
| →1-2¶ | 68 (7.3) | 267 (11.5) |
| →3-6¶ | 355 (38.1) | 922 (39.8) |
| →7 and more☐ | 448 (48.0) | 974 (42.1) |
| Mother listens to radio☑ |  |  |
| →Almost everyday☑ | 60 (6.2) | 143 (6.4) |
| →Not at all/less than once/at least once a week☐ | 873 (93.8) | 2,172 (93.6) |
| Mother reads newspaper☑ |  |  |
| →Almost everyday☑ | 109 (11.8) | 536 (23.2) |
| →Not at all/at least once a week☐ | 823 (88.2) | 1,779 (76.8) |
| Mother listens to television☑ |  |  |
| →Almost everyday | 672 (72.0) | 1,926 (83.2) |
| →Not at all/at least once a week | 261 (28.0) | 389 (16.8) |
| Wealth quintile☑ |  |  |
| →Poorest | 369 (39.6) | 380 (16.4) |
| →Poorer | 259 (27.7) | 476 (20.6) |
| →Middle | 151 (16.2) | 459 (19.8) |
| →Richer | 98 (10.5) | 511 (22.1) |
| →Richest☐ | 55 (5.9) | 488 (21.1) |
| *Community level characteristics¶¶* |  |  |
| Class¶¶ |  |  |
| →Upper class¶¶ | 349 (48.6) | 700 (31.0) |
| →Scheduled caste¶¶ | 279 (30.8) | 451 (19.9) |
| →Scheduled tribe¶¶ | 32 (3.6) | 56 (2.5) |
| →Other marginalised class☐ | 210 (23.2) | 934 (41.4) |
| State¶¶ |  |  |
| →Delhi | 99 (10.6) | 949 (41.0) |
| →Maharashtra | 625 (67.0) | 286 (12.4) |
| →Tamil Nadu | 25 (2.8) | 192 (8.3) |
| →Uttar Pradesh | 52 (5.6) | 173 (7.5) |
| →West Bengal | 43 (4.6) | 166 (7.2) |
| →Telangana | 33 (3.5) | 372 (16.1) |
| →Madhya Pradesh☐ | 55 (5.9) | 177 (7.7) |
| *Policy level¶¶* |  |  |
| Place of birth¶¶ |  |  |
| →Home | 55 (6.0) | 177 (7.7) |
| →Health facility□ | 877 (94.1) | 2134 (92.3) |

*Note:* N=number of all participants n=number of participants in each subset, %=percentages

### Community-level characteristics of the participants

In the urban slum areas, the participants were more in the upper class (48.6%) than the other societal classes (30.8% in the scheduled castes, 23.2% in the marginalized class and 3.6% in the scheduled tribes). Whereas in the urban non-slum, most of the participants were in the marginalized class (41.4%) compared to the upper class (31.0%). 67.0% of the participants living in the urban slum were from Maharashtra, and urban non-slum participants were mostly from Delhi (41.0%) compared to the other six states

### Prevalence of breastfeeding practices by area of residence: urban slum and non-slum areas in India

The prevalence of EBFI within an hour was higher in the urban slum (50.4%, 95% CI: 47.2-53.7) than in the urban non-slum areas (37.4%, 95% CI: 35.4-39.4) (Fig 1), but the difference was not statistically significant (p=0.768). EBF was significantly higher in urban non-slum (55.8%, 95% CI: 51.5-60.0) than in urban slum areas (50.0%, 95% CI: 42.5-57.7) (p=0.004).

**Fig 1.**
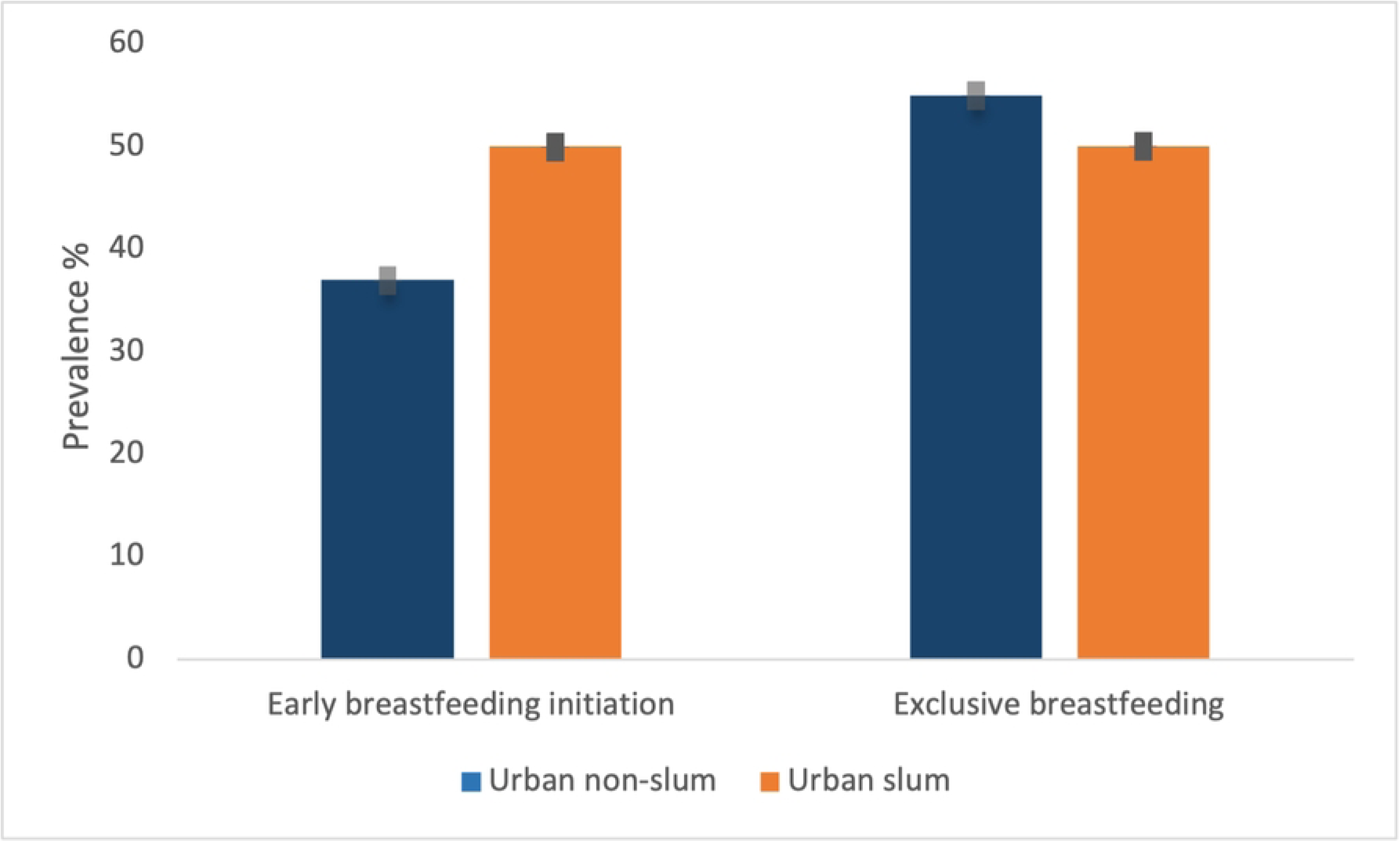
Prevalence of breastfeeding practices stratified by urban slum and non-slum areas in India.

Early initiation of breastfeeding varied considerably across states and between urban slum and non-slum areas. Tamil Nadu (67.4%), Telangana (63.7%) and Maharashtra (58.3%) reported the highest EIBF rates among slum populations, while West Bengal (60.9%) and Maharashtra (61.1%) led among non-slum populations. Notably, Telangana showed a substantial slum–non-slum gap (63.7% vs. 39.7%), while Uttar Pradesh recorded the lowest EIBF rates in both settings (<9% (Table 2). The prevalence of EBF across the states was significantly higher in the urban non-slum areas than the urban slum areas, except in Madhya Pradesh, Uttar Pradesh, and Delhi

**Table 2.** State prevalence of early breastfeeding initiation practices of 7 states stratified by urban slum and non-slum areas in India.

| States | Early breastfeeding initiation |  |
| --- | --- | --- |
|  | Urban slum<br>n(%) | Urban non-slum<br>n(%) |
| →Delhi | 35 (38.4) | 288(31.5) |
| →Maharashtra | 353 (58.3) | 169(61.1) |
| →Tamil Nadu | 16 (67.4) | 78(45.3) |
| →Uttar Pradesh | 2 (4.71) | 14(8.3) |
| →West Bengal | 10 (23.6) | 98(60.9) |
| →Telangana | 20.1 (63.7) | 147(39.7) |
| →Madhya Pradesh□ | 17 (31.7) | 42(24.2) |

Exclusive breastfeeding rates showed wide variability across states and settlement types. Among urban slums, Madhya Pradesh reported the highest EBF prevalence (95.6%), while Tamil Nadu reported none. In non-slum areas, Telangana (75.6%) and Maharashtra (74.5%) had the highest rates. Uttar Pradesh had the lowest EBF levels in both slum (18.3%) and non-slum (15.7%) areas (Table 3).

**Table 3.** State prevalence of exclusive breastfeeding practices (<6months old) stratified by urban slum and non-slum areas in India.

| States | Exclusive breastfeeding |  |
| --- | --- | --- |
|  | Urban slum<br>n(%) | Urban non-slum<br>n(%) |
| →Delhi | 12 (52.2) | 96 (49.3) |
| →Maharashtra | 49 (54.1) | 46 (74.5) |
| →Tamil Nadu | 0 (0) | 38 (62.1) |
| →Uttar Pradesh | 3 (18.3) | 7 (15.7) |
| →West Bengal | 3 (38.1) | 15 (48.9) |
| →Telangana | 3 (63.7) | 70 (75.6) |
| →Madhya Pradesh<sup>a</sup> | 12 (95.6) | 19 (52.8) |

### Individual-level correlates of breastfeeding practices

At the individual level in urban slum areas, mothers who reported one to two antenatal care visits were less likely to practice exclusive breastfeeding (adjusted OR: 0.04, 95% CI: 0.00-0.72) compared to mothers who had no antenatal visit (Table 5).

**Table 4.**
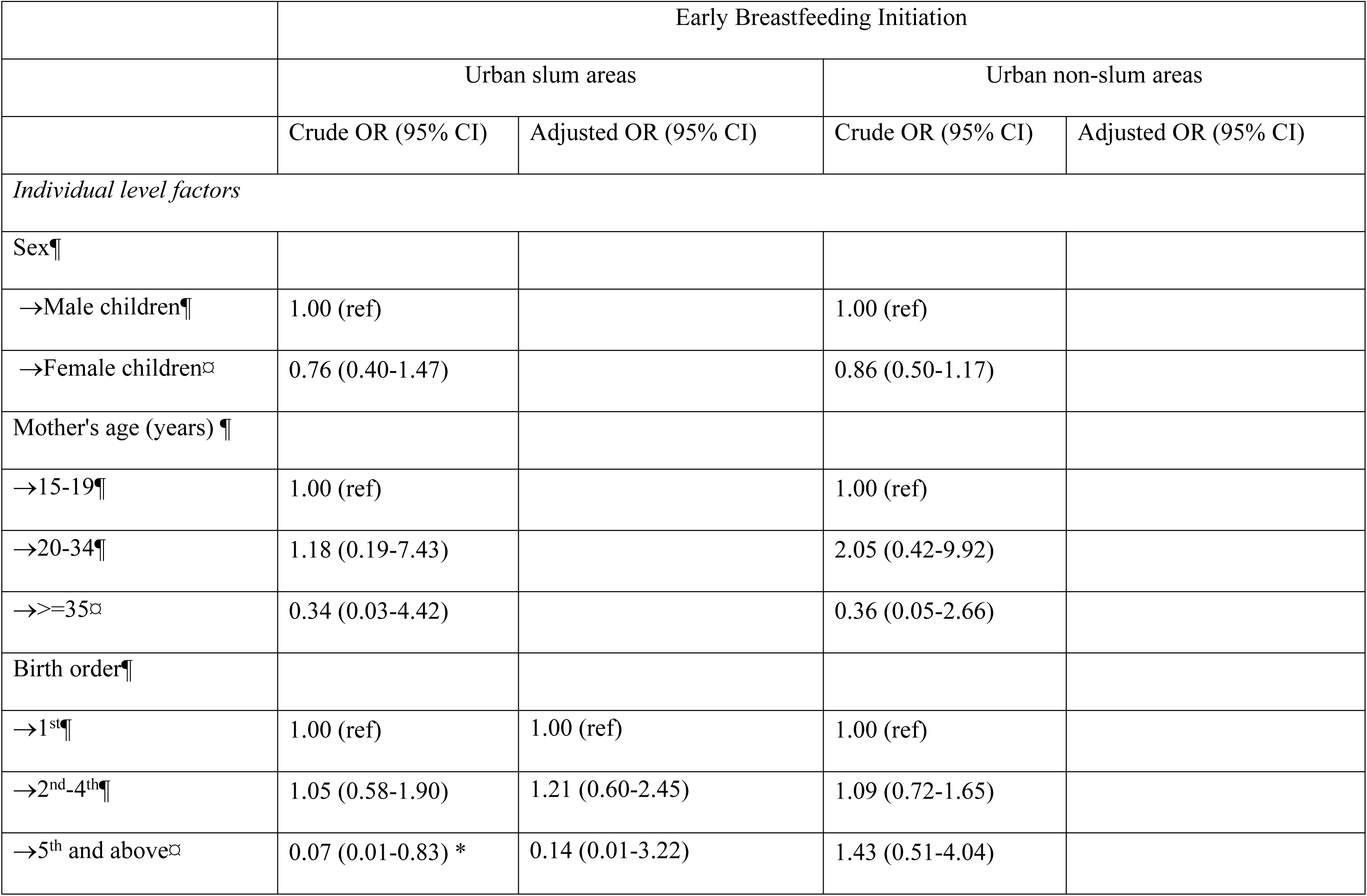
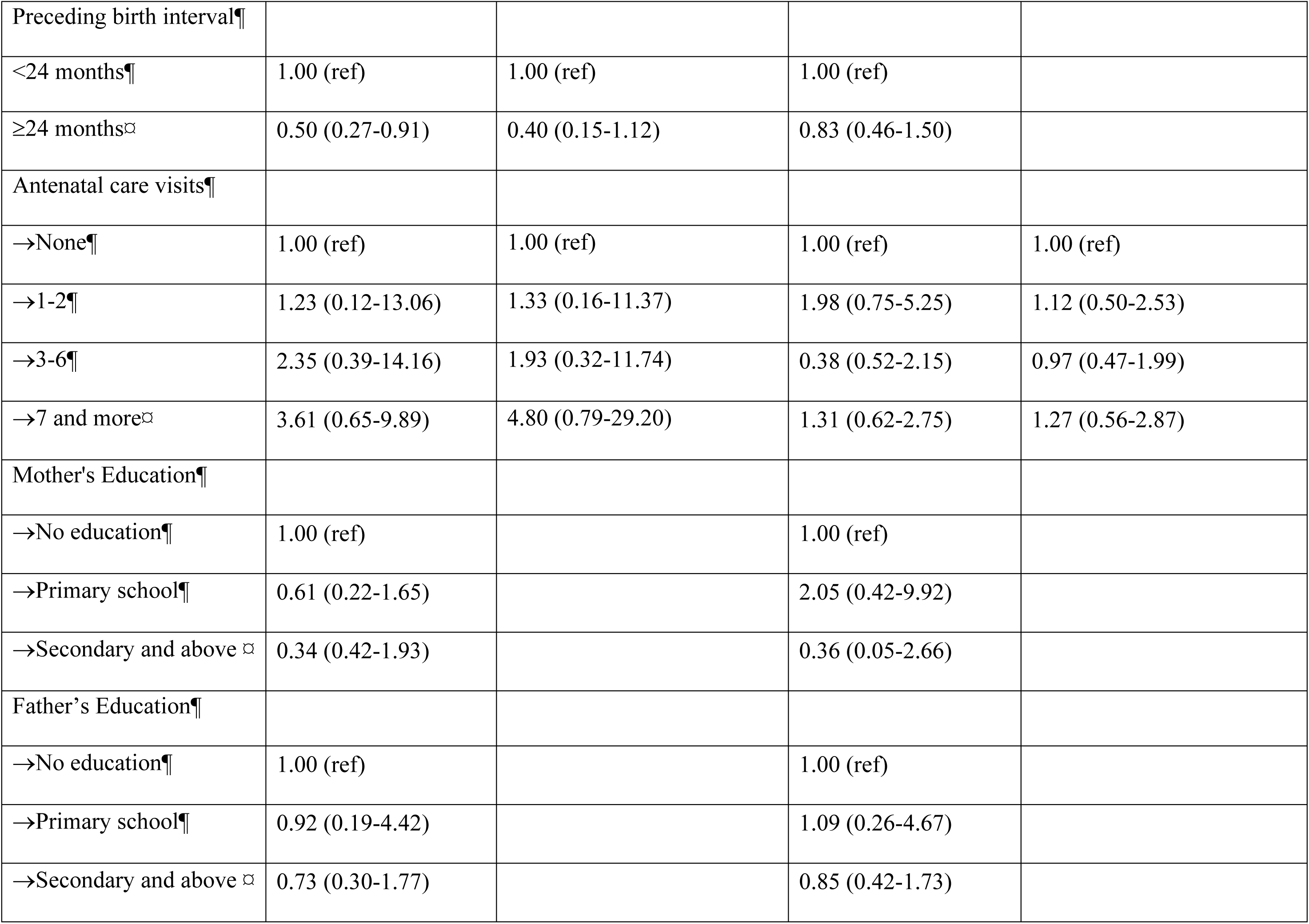
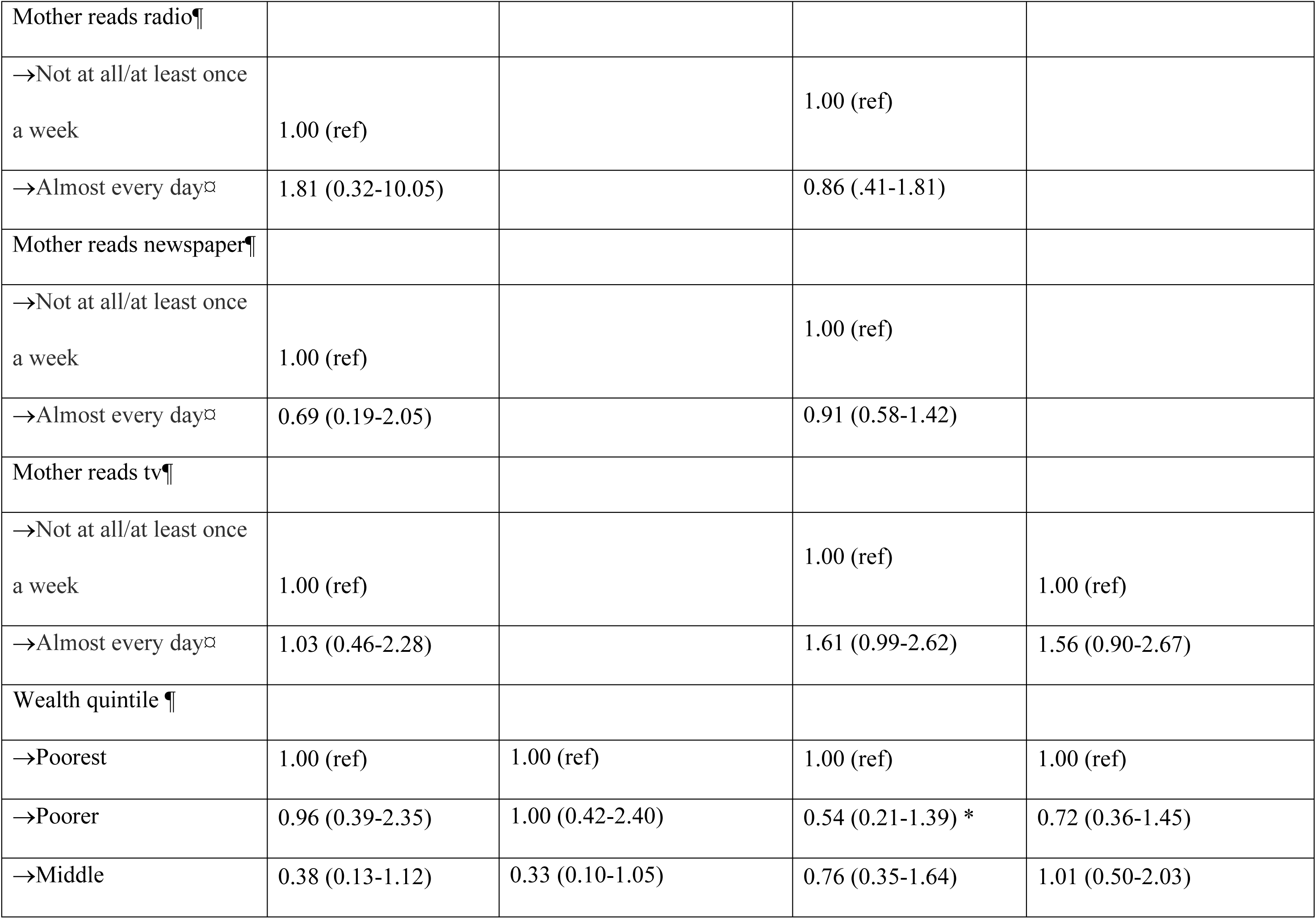
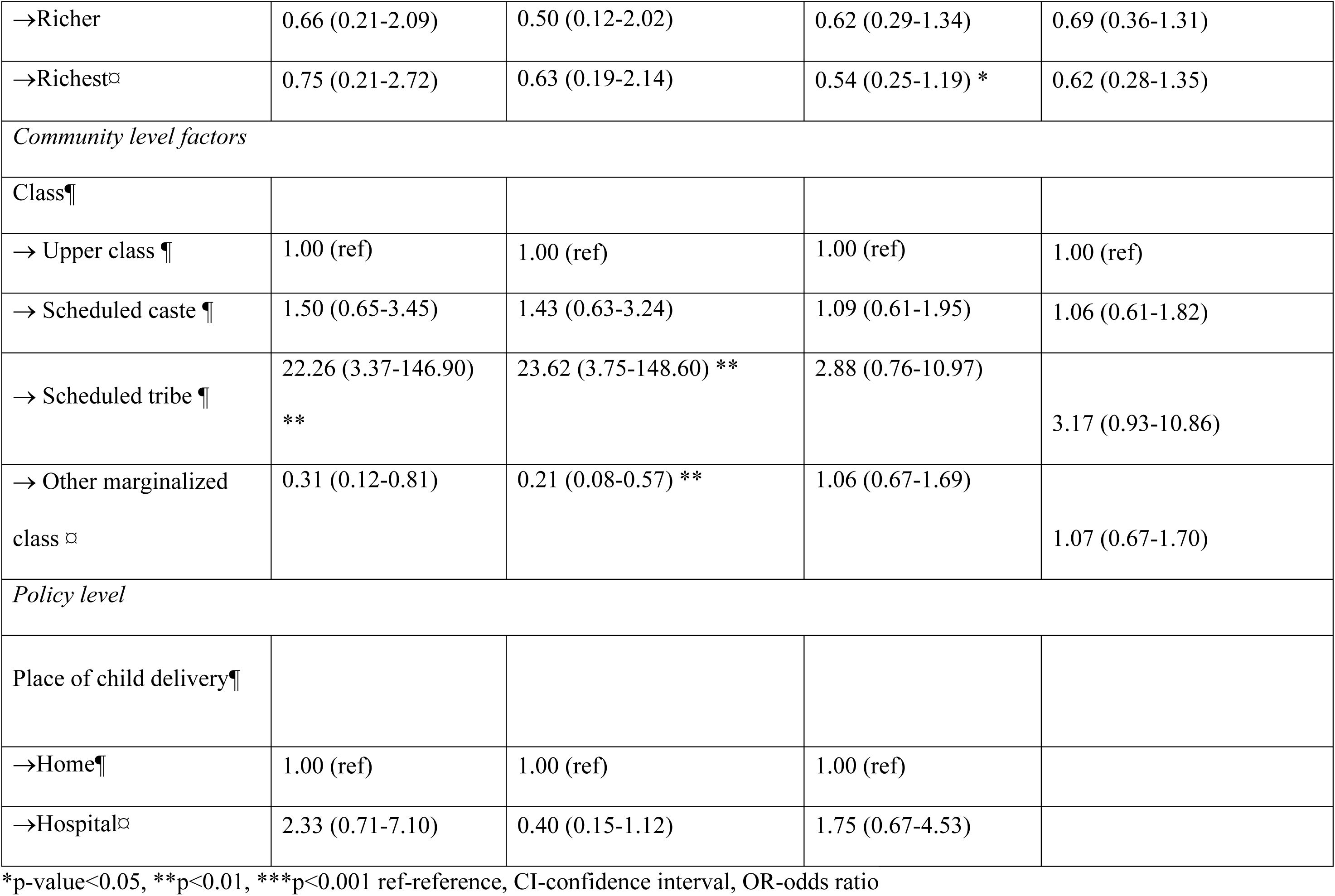
Correlates of Early Breastfeeding Initiation Practices stratified by Urban slum and non-slum areas in India.

**Table 5.**
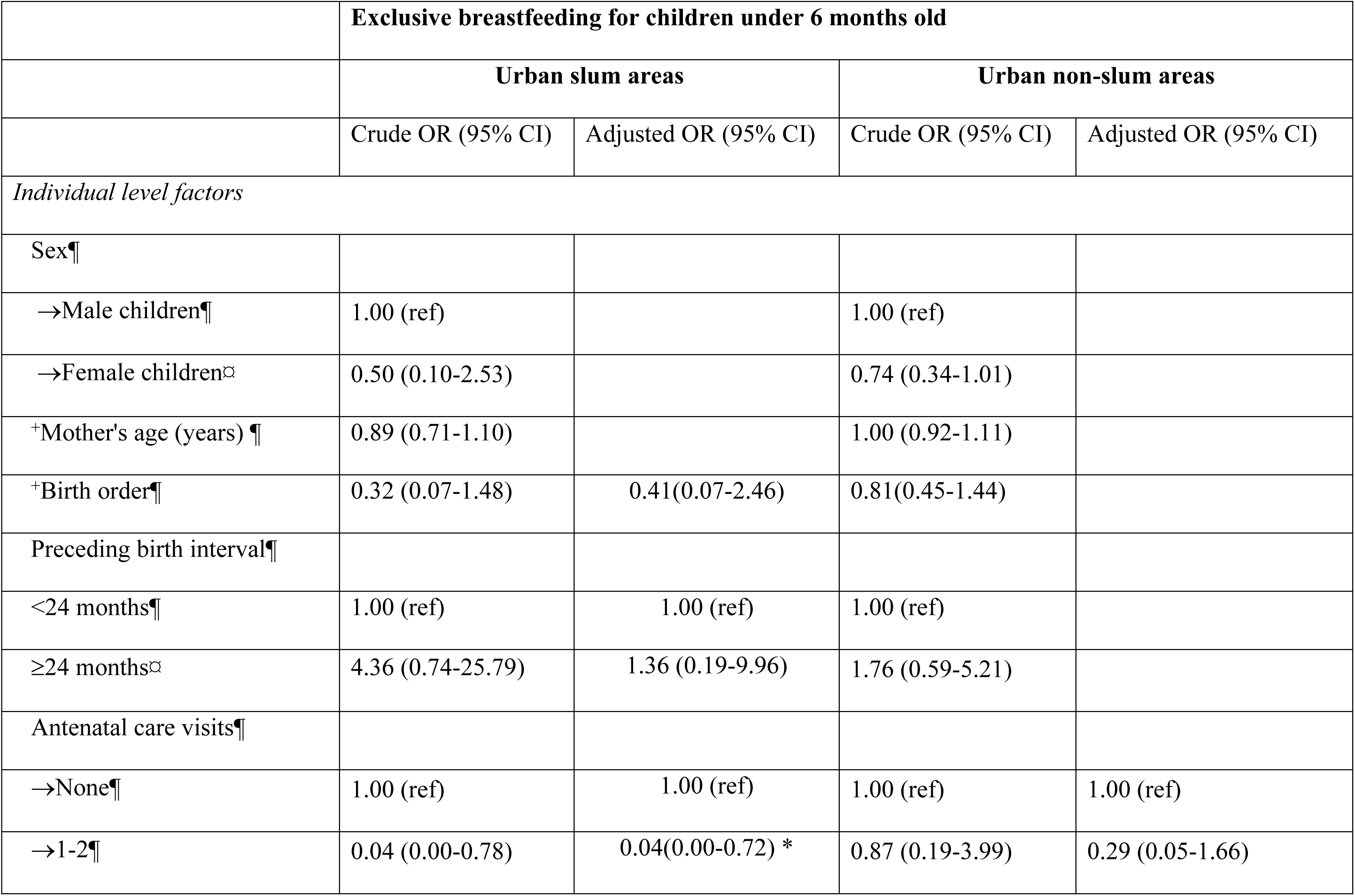
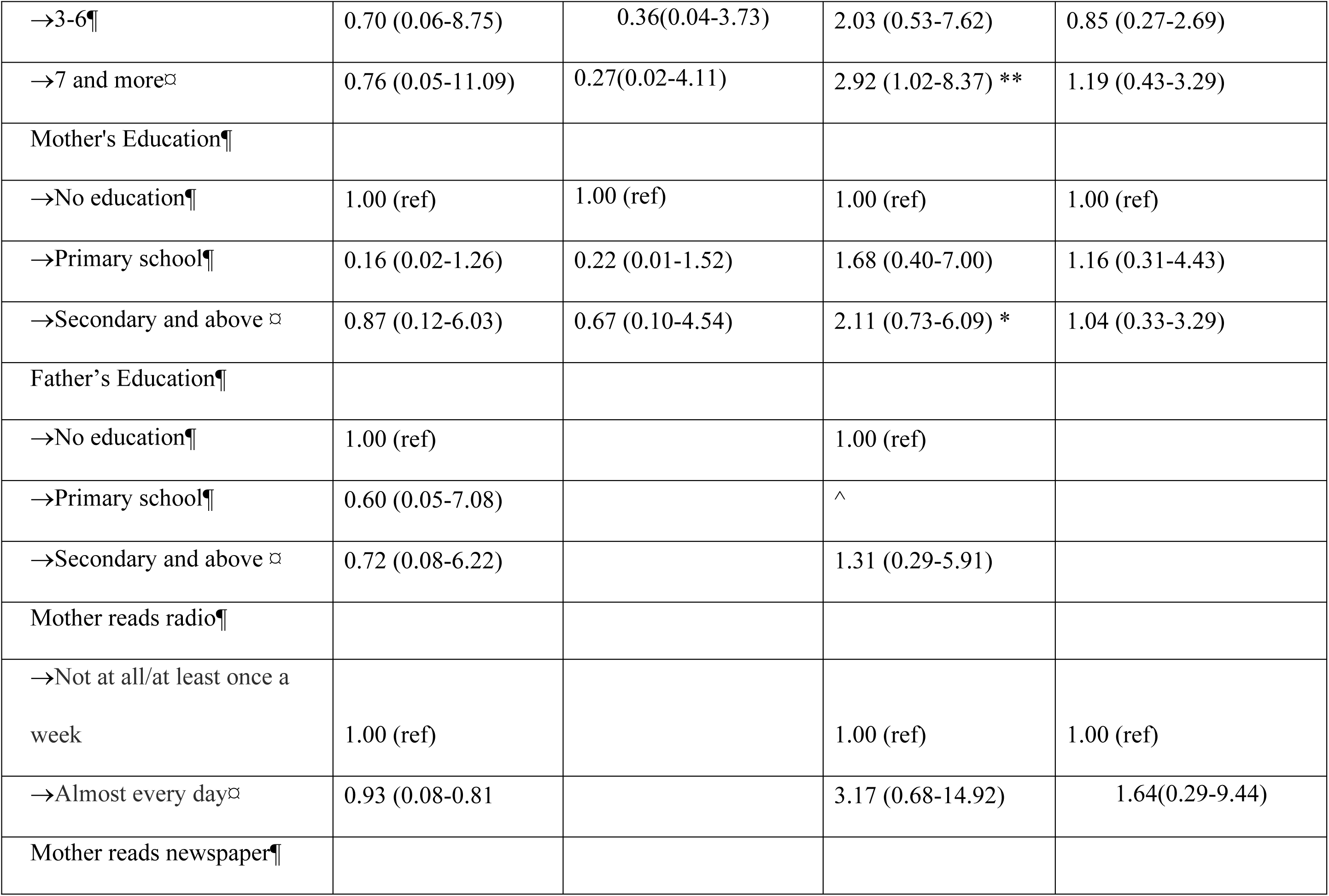
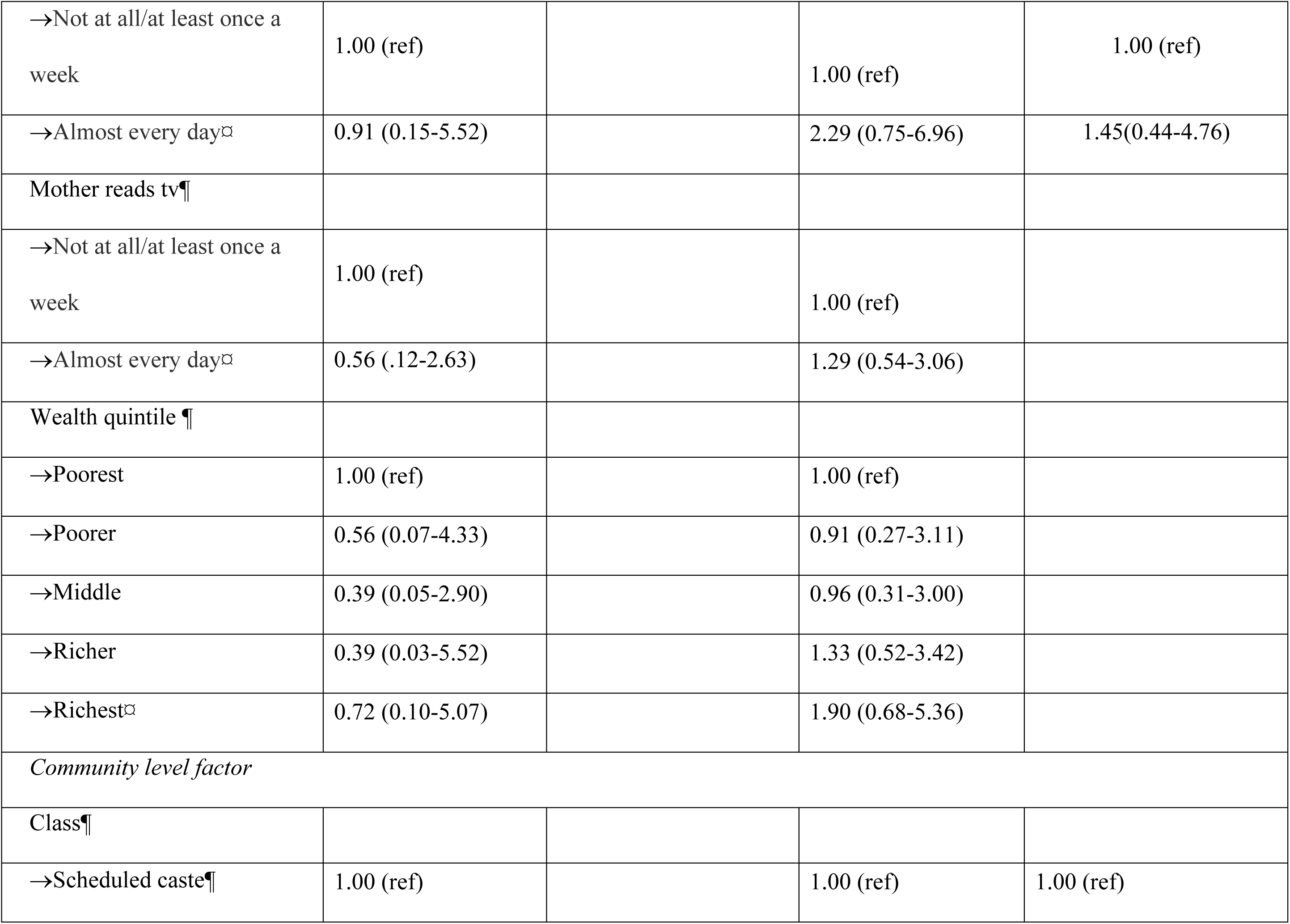
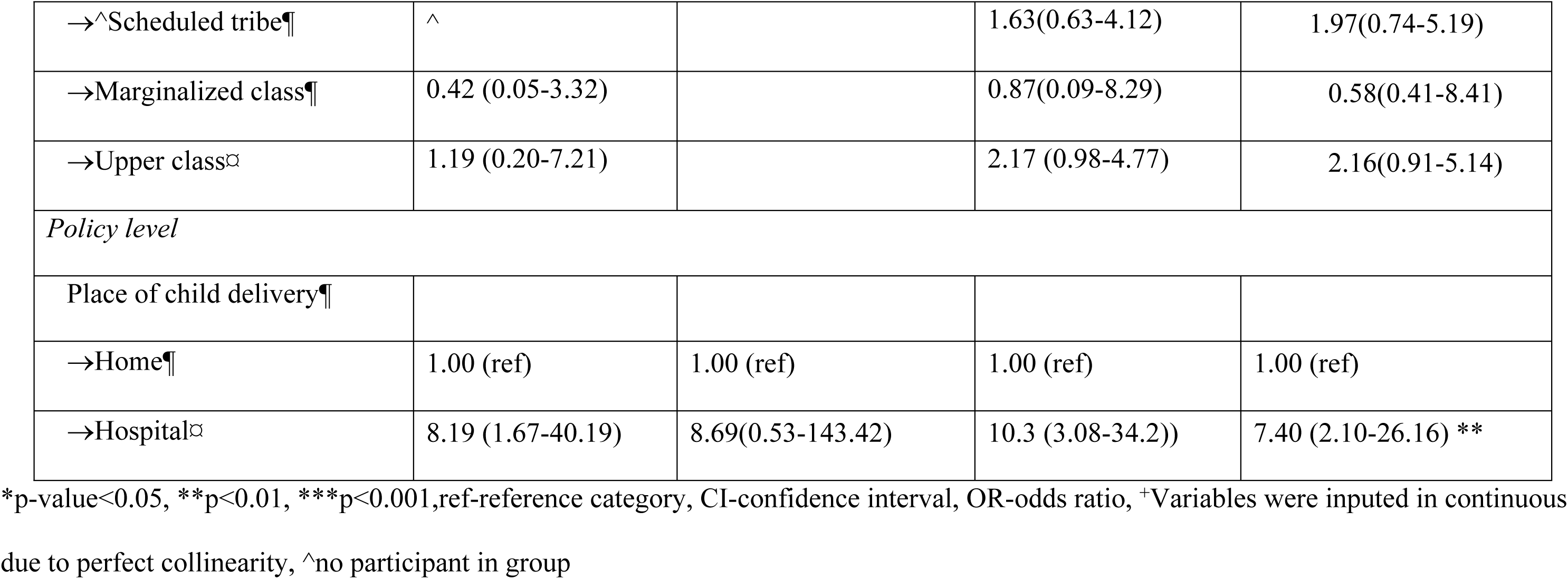
Correlates of exclusive breastfeeding for children aged less than 6 months old stratified by urban-slum and urban-non slum in India.

### Community-level correlates of breastfeeding practices

In the slum areas, our final model showed that Scheduled Tribes were associated with increased odds of practicing EBFI (adjusted OR= 23.62, 95%CI:3.75-148.60), whereas belonging to the Other Marginalized Class was associated with less likelihood of practicing EBFI (adjusted OR=0.21, 0.08-0.57) (Table 4). While in the non-slum areas, no significant associations were identified for EIBF, and EBF had no significant associations in either area.

### Policy-level correlates of breastfeeding practices

In the adjusted model for non-slum areas (adjusted OR: 7.40, 95%CI: 2.09-26.16) children born in the health facilities were more likely to be exclusively breastfed than children not born in the health facilities. However, no significant associations were observed for EBFI in either area at the policy level (Table 3).

## Discussion

In our study, only half of the mothers in the slum initiated breastfeeding within an hour of birth, which is consistent with a study in East Delhi (28). We also found that less than half (37%) of the mothers in our study practiced EBFI in the non-slum areas, which is below the national average of 41% (9). EBF was lower in urban slums than in urban non-slum areas (55%). However, other studies reported lower rates of EBF in slums, and this may be explained by the inconsistent definitions used for EBF indicators(14–16). Based on the WHO definition, which we applied, the age band for assessing EBF is 0-5 months, but other studies included 6-month-old babies who may have commenced complementary feeding, hence their underestimation (14–16).

Across the seven states where slums were sampled, EBFI was generally higher in urban slums compared to non-slum areas in Delhi, Tamil Nadu, Madhya Pradesh, and Telangana, whereas non-slum areas showed higher EIBF in Maharashtra, West Bengal, and Uttar Pradesh. EBF patterns varied, with slum areas reporting higher rates in Madhya Pradesh and slightly in Uttar Pradesh, but notably lower or absent in Tamil Nadu, Maharashtra, West Bengal, and Telangana. EBF rates in Delhi were comparable across both settings. These findings highlight substantial geographic and contextual disparities in breastfeeding practices between urban slum and non-slum populations. According to the global collective targets for 2030, which is 70% for EBFI and 70% for EBF, there is an urgent need to amplify efforts across the seven states, especially Uttar Pradesh and Tamil Nadu (29).

Our study revealed correlates of breastfeeding practices at the individual, community, and policy levels. At the individual level, attending one to two antenatal care visits (ANC) in the urban slum was associated with less practice of exclusively breastfeed compared to no antenatal visits. Notably, mothers with higher numbers of ANC visits did not show a significantly increased likelihood of EBF either. This pattern suggests that attending ANC, regardless of frequency, may not be sufficient to promote optimal breastfeeding practices. This may reflect broader institutional or systemic challenges negatively affecting maternal health behaviours, including EBF (30).

At the community level, being in the marginalized class was associated with lower odds of EIBF, in contrast to belonging to the scheduled tribes, which was associated with increased odds of EBF. These findings mirror those of another study of the East and North-East regions of India (22). A possible explanation for this is the practice of discarding colostrum, which is prevalent amongst the socioeconomically disadvantaged class. Our study did not collect information on the discard of colostrum, therefore, we are unable to attribute our observation to this practice (31). Future research will be required to determine the precise factors responsible for breastfeeding practice inequity between marginalized and other classes in India.

At the policy level, in non-slum areas, children born at the hospital were more likely to be exclusively breastfed compared to children born in other settings outside the health facilities. A study conducted in Nagpur slum in India reported the same findings, and this is possibly because health personnel in the facilities are likely to promote breastfeeding practices, and mothers may benefit directly from counselling received from these health personnel when being attended to (14,32). With over 90% of mothers in the urban slums and urban non-slum areas delivering children at health facilities, the Mother Absolute Affection program, which is a Baby Friendly Hospital Initiative by the Ministry of Health and Family Welfare of the Government of India to increase breastfeeding rates, needs to be further strengthened to ensure quality, effective implementation and inclusion of mothers in the urban slum areas. For Uttar Pradesh (slum and non-slum areas) and Tamil Nadu (slum), where EBFI and EBF were below other states, their health facilities should be monitored for adherence to EBFI, and health workers should be well-trained to support mothers on EBFI and EBF. As reported in India and Bangladesh where community health workers who provided essential newborn care contributed to 54% and 34% reduction in neonatal deaths respectively, collaboration with community health workers should be encouraged to ensure widespread diffusion of nutrition messaging for ease of implementation.(33,34).

The primary limitation of this study is the use of a 24-hour recall assessing children’s feeding practices, which may have been subject to recall bias. To minimize this bias, the analysis focused on the last-born children living with their mothers. Another limitation is the potential for social desirability bias in responses regarding feeding practices, particularly when incorrect practices are disapproved of within the community, possibly leading to an overestimation of correct feeding practices. Additionally, over 80% of data on mothers’ occupations were missing, preventing further analysis of this variable. Lastly, the sample size for assessing exclusive breastfeeding (EBF) among children under 6 months in the slum was small.

Despite these limitations, our study has strong external validity and potential implications for research, policy, and practice. To our knowledge, this is the first study to conduct a comparative analysis of breastfeeding practices between urban slums and urban non-slum areas in India using DHS data. Our data represents six geographical regions of India (Southern, Western, Eastern, Central, Northern, and North-eastern) collected with standardized questionnaires, which have been utilized for over three survey cycles for India (1998–2015), and our outcomes which were assessed based on WHO standard criteria strengthens the accuracy of our results. Due to their wide geographical coverage, we believe our findings are generalizable to all urban slums and urban non-slum areas in India and, therefore, applicable to the overall improvement of breastfeeding practices in the country.

## Conclusion

Breastfeeding practices were suboptimal in both urban slum and non-slum areas in India. The prevalence of EIBF was higher in the urban slum than in urban non-slum areas, while EBF prevalence was higher in the urban non-slum than urban slum areas. Individual, community, and policy-level factors were associated with breastfeeding practices in India. We recommend policy reforms to support the overall living conditions of children living in the slums and engage ethnic minority support groups to improve breastfeeding practices in India. A long-term recommendation would be to address poverty and increase access to quality healthcare for slum dwellers. We also urge future studies to explore the mechanisms behind the observed associations in our studies using qualitative methods, as this would strengthen health improvement interventions at individual, community, and policy levels. Lastly, with no associations observed in the non-slum areas for EBFI, future research should examine other contextual factors like insecurity, marginalization, policies, communicable diseases, and environmental factors like climate-induced droughts and flooding that may play a role in shaping breastfeeding practice.

## Data Availability

The Demographic and Health Surveys (DHS) data are publicly available data that can be downloaded from the DHS Program’s website (URL: https://www.dhsprogram.com/). The specific data set used by the authors was the 2015-16 India DHS survey.

## Acknowledgments

The authors thank Rishi Caleyachetty for his unreserved guidance, support, and follow-up for this study.

